# SARS-CoV-2 variants of concern Alpha and Delta show increased viral load in saliva

**DOI:** 10.1101/2022.02.10.22270797

**Authors:** Kylie L. King, Stevin Wilson, Justin M. Napolitano, Keegan J. Sell, Lior Rennert, Christopher L. Parkinson, Delphine Dean

## Abstract

**Background:** Higher viral loads in SARS-CoV-2 infections may be linked to more rapid spread of emerging variants of concern (VOC). Rapid detection and isolation of cases with highest viral loads, even in pre- or asymptomatic individuals, is essential for the mitigation of community outbreaks.

**Methods and Findings:** In this study, we analyze Ct values from 1297 SARS-CoV-2 positive patient saliva samples collected at the Clemson University testing lab in upstate South Carolina. Samples were identified as positive using RT-qPCR, and clade information was determined via whole genome sequencing at nearby commercial labs. We also obtained patient-reported information on symptoms and exposures at the time of testing. The lowest Ct values were observed among those infected with Delta (median: 22.61, IQR: 16.72-28.51), followed by Alpha (23.93, 18.36-28.49), Gamma (24.74, 18.84-30.64), and the more historic clade 20G (25.21, 20.50-29.916). There was a statistically significant difference in Ct value between Delta and all other clades (all p.adj<0.01), as well as between Alpha and 20G (p.adj<0.05). Additionally, pre- or asymptomatic patients (n=1093) showed the same statistical differences between Delta and all other clades (all p.adj<0.01); however, symptomatic patients (n=167) did not show any significant differences between clades. Our weekly testing strategy ensures that cases are caught earlier in the infection cycle, often before symptoms are present, reducing this sample size in our population.

**Conclusions:** COVID-19 variants Alpha and Delta have substantially higher viral loads in saliva compared to more historic clades. This trend is especially observed in individuals who are pre- or asymptomatic, which provides evidence supporting higher transmissibility and more rapid spread of emerging variants. Understanding the viral load of variants spreading within a community can inform public policy and clinical decision making.

## Introduction

The United States confirmed its first positive SARS-CoV-2 case on January 21, 2020 [1]. As of December 1, 2021, there have been over 265 million cases globally and 48 million in the United States alone. Following its emergence in December 2020, clade 21A (classified as the Delta variant) spread rapidly across the globe. On May 29, 2021, the CDC reported that 7.3% of new cases in the U.S.A. were identified as Delta, and 65.4% of cases were clade 20I (Alpha). By August 28, 99.1% of reported cases were Delta [1]. This rapid shift may be attributed to key mutations that increase transmissibility, due in part to a higher viral load.

In early 2021, the Alpha variant spread rapidly due to the N501Y mutation in the S protein which enhances its affinity for angiotensin-converting enzyme 2 (ACE2), the cellular receptor that facilitates viral entry [2]. The Delta variant lacks this mutation but carries several mutations within the S protein; specifically, L452R, T478K, and P681R, which confer resistance to monoclonal antibody treatments [3]. The L452R and T478K mutations may also increase transmissibility of the virus by stabilizing the ACE2-receptor binding domain (RBD) complex [3]. Another mutation within the N protein, R203M, increases viral mRNA delivery and expression, allowing the Delta variant to produce >50-fold more viral particles [4]. These mutations may improve host cell binding affinity, as well as increase viral production, and may contribute to the rapid global spread of this variant.

Most studies of SARS-CoV-2 viral loads used the nasal or nasopharyngeal (NP) swab sample collection method [5-7]. The viral load in saliva and oral swab samples has been correlated with COVID-19 symptoms and transmissibility, and have been suggested to be similarly or slightly more sensitive than nasal swabs early in the infection cycle [8-14]. Low Ct values are associated with high viral load and increased transmissibility, primarily due to viral presence in saliva droplets that facilitate spread when infected individuals are in proximity [7, 15-17]. Saliva has been shown to be an accurate diagnostic tool, yielding comparable Ct values to NP swabs while decreasing cost per test, discomfort to patients, and risk of transmission to healthcare workers during collection [9-10,13,18].

## Methods

### Sample Collection

Ethical review for this study was obtained by the Institutional Review Board of Clemson University. This is a retrospective study on archived de-identified samples and data. The samples and data sets were stripped of patient identifiers prior to any SARS-CoV-2 sequencing and data analysis.

To evaluate the relative viral load of the variants of concern (VOC) found in upstate South Carolina (Alpha, Gamma, and Delta), we compared the Ct values from saliva samples from the SARS-CoV-2 testing lab at Clemson University, which also provides free testing for the surrounding community [19-20]. University surveillance testing is mandatory for students and employees on a weekly or bi-weekly schedule regardless of vaccination status [21]. The study population includes all university students and employees, as well as members of the surrounding community that tested positive between January and November 2021. Samples were labeled as “symptomatic” if the patient self-reported symptoms at the time of collection, or “exposed” if they reported recent viral exposure. All other samples were considered “surveillance”. Only one positive test was included for each patient; any subsequent tests were excluded from our analysis.

### SARS-CoV-2 Identification and Sequencing

SARS-CoV-2 positive saliva samples were identified using the TigerSaliva multiplex RT-qPCR testing method, which targets the N gene [19]. The TigerSaliva diagnostic assay is a version of the EUA-approved SalivaDirect protocol [11] that utilizes open-source sample handlers (Opentrons OT-2) and standard thermocycler (Bio-Rad CFX 384) systems. Briefly, 1mL of saliva is collected from patients in standard 50mL conical tubes. The saliva is heated to 95°C for 30 minutes before 2μL are loaded into test plates with enzyme mix, primers, and probes. The assay measures the N1 sequence of SARS-CoV-2 and Hs_RPP30 (human control gene). The N-gene of SARS-Cov-2 is a single-copy gene, thus 1 copy of the N-gene is equivalent to 1 copy of the virus. This protocol was found to have a 90% sensitivity and 99% specificity when compared to paired NP swabs [19]. It was determined by standard curve that a Ct of 33 was equivalent to 1 viral copy per microliter (cpu) of saliva (SFig 1) and was therefore used as the cutoff for positivity. Note that samples with viral loads less than 1cpu can be measured, however they are not considered positive as per diagnostic protocols [11,19]. Samples were run in duplicate, and the average Ct value from both replicates was used for this analysis.

Heat-treated saliva samples were commercially sequenced (Premier Medical Sciences, Greenville, SC; Labcorp, Durham, NC) using the ARTIC protocol. Briefly, RNA was extracted from saliva samples via MagBind Viral RNA Kit (Omega Biotek, Norcross, GA) and recovered SARS-CoV-2 RNA was quantified via Logix Smart COVID-19 assay (Co-Diagnostics, Salt Lake City, UT). Samples with sufficient RNA were sequenced on the Illumina NovaSeq 6000 or NextSeq500/550 according to manufacturer’s protocols. Sequences were assembled and analyzed using nf-core/viralrecon v.2.2 [22]. Sequence data was uploaded to SC DHEC, GenBank, and GISAID (see Supplementary Data). These databases have requirements regarding the number of ambiguous nucleotides allowed in the consensus sequence. Some of the samples in this analysis exceeded this threshold, which prevented database upload, but all had sufficient information to confidently assign clade by Pangolin and Nextclade [23-24].

### Statistical Analysis

Ct values among VOCs were compared: 20I (Alpha), 21A (Delta), 20G, and 20J (Gamma, V3) [23]. Due to low prevalence in the Upstate South Carolina community, 20H (Beta) samples (n=8) were excluded from analysis. To maintain phylogenetic independence, we only compare Ct values for variants at branch tips within the NextClade phylogeny [24]. Therefore, the four Nextclades we compare do not have parent-offspring relation. Clades 20A (n=65) and 20B (n=29) were excluded from this analysis. Statistical analyses were performed in R using Kruskal-Wallis test followed by Dunn’s test of multiple comparisons.

## Results and Discussion

We first determined the clade composition in our community from positive samples collected between January and December 2021 (Fig 1). We only sequenced samples that tested positive via the TigerSaliva assay (Ct<33). We did notice that samples within the higher Ct range (28-33) had more regions with ambiguous nucleotides and were therefore less likely to have received a clade assignment via Nextclade. From January to July, we sequenced all positive samples stored from the lab. Due to the increase in positive samples during the Delta surge, we sequenced a statistical sampling of positives (approximately 15%) to ensure accurate representation of our community demographics.

**Fig 1.**
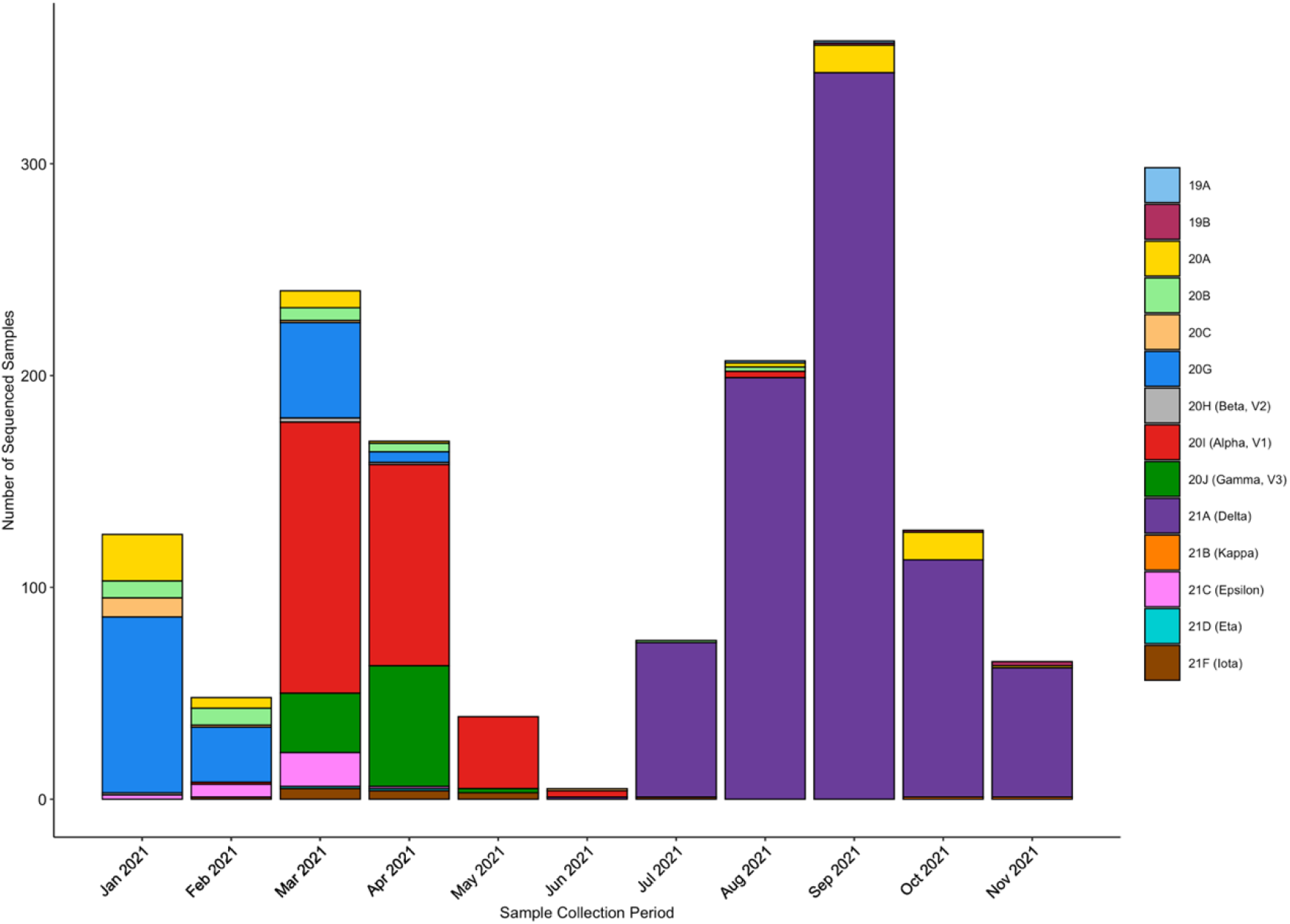
Clade composition of samples run in the REDDI Lab from January to December 2021. Clade determination was made via whole genome sequencing. There were few positive samples between May and June 2021 due to the university summer break.

In this study there were no differences in Ct values based on vaccination status. It is important to note that the percentage of vaccinated individuals prior to June 2021 is very low, particularly in those under 40 years old, as the majority were not eligible for vaccination until mid-April. By August 2021, approximately 40-45% of adults in our region were vaccinated; very few minors (12-18 years old) were vaccinated, and vaccines were not available for children younger than 12. There were no observable trends in Ct values between a particular variant and any other demographic factors considered: age, gender, etc. However, there was a significant difference in patient age between clades; the average ages of patients infected with the Delta and Gamma variants were significantly younger than the Alpha or 20G variants (p<0.001, see Supplementary Data). The Gamma variant emerged in our community following the university Spring break, likely due to the travel of undergraduate students. The Delta surge was notable in that it was characterized with large outbreaks in K-12 schools, which were open to in-person instruction in early August 2021. In the previous Spring 2021 semester schools were open with multiple mitigation measures in place to prevent outbreaks (e.g., hybrid instructions, social distancing, masking) and there were very few cases of COVID-19 in children [25]. But, in the Fall 2021 academic term K-12 schools in South Carolina were prohibited from imposing mask mandates or switching to hybrid instruction due to state legislation passed during summer 2021 [26].

SARS-CoV-2 positive samples showed a significant difference between Delta (median: 22.61, IQR: 16.72-28.51) and all other clades [Alpha: 23.93 (18.36-28.49), Gamma: 24.74 (18.84-30.64), 20G: 25.21 (20.50-29.916)] (Fig 2). When only surveillance samples were considered (Fig 2B), the same trend was observed with Delta (median: 22.56, IQR: 16.67-28.45) having a significantly lower median Ct from other clades [Alpha: 23.81 (18.51-29.11), Gamma: 24.69 (18.84-30.54), 20G: 25.75 (21.53-29.98)]. Additionally, both groups showed a significant difference in Ct values between Alpha and 20G.

**Figure 2:**
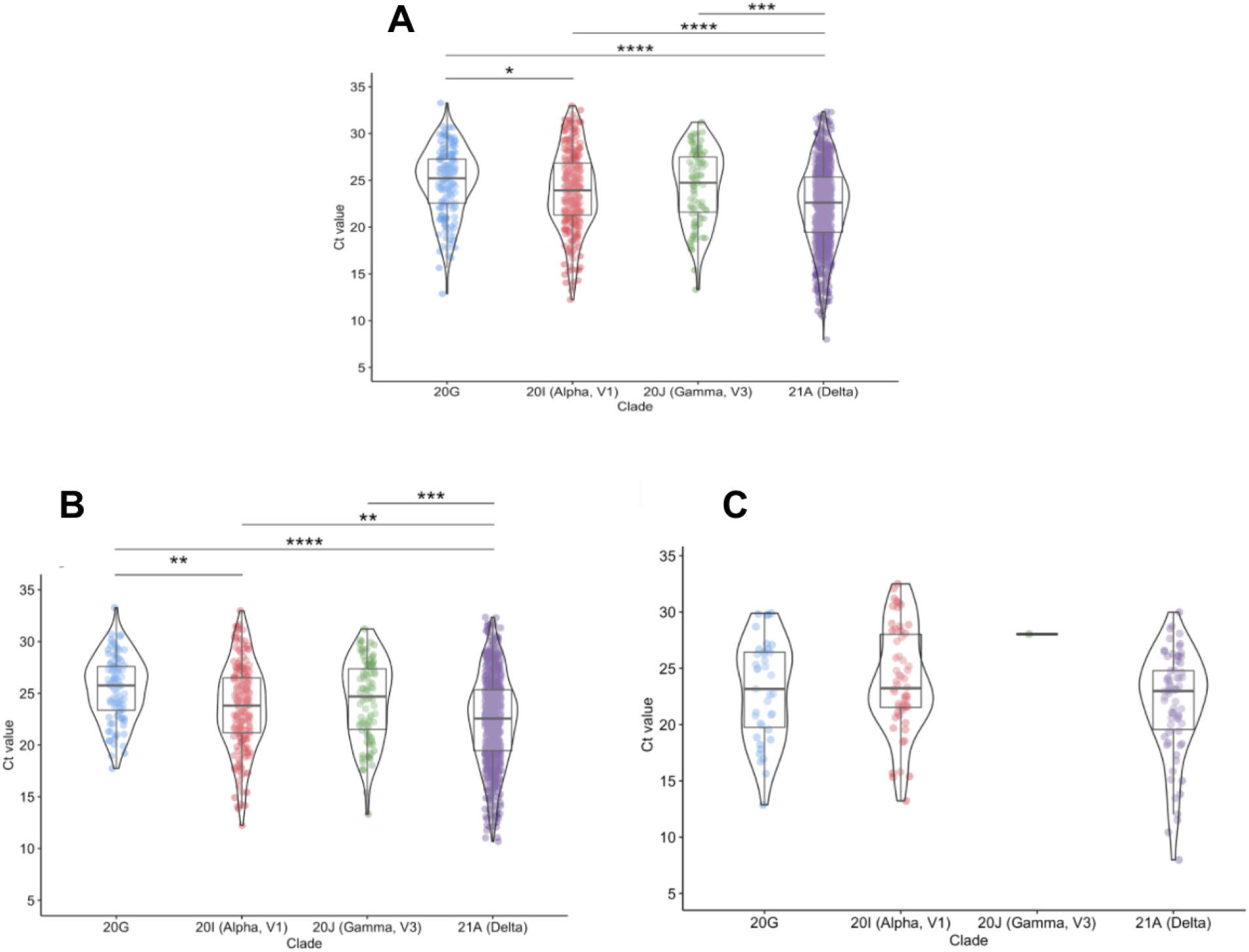
N1 Ct values of common clades in saliva. We analyzed the Ct values from a total of 1297 SARS-CoV-2 positive saliva samples, using the N gene target. **2A: Comparison of all samples**. Delta (n=787) showed a statistically significant difference in Ct value when compared to 20G (n=159), Alpha (n=258), and Gamma (n=87). **2B: Comparison of surveillance samples**. When only surveillance samples were considered, the same trends were observed, showing a significant difference between Delta (n=691) and all other clades (20G: n=95, Alpha: n=181, Gamma: n=86). Both groups also showed a significant difference when comparing Alpha and 20G. **2C. Comparison of symptomatic samples**. There were no significant differences in Ct values observed among symptomatic samples for Delta (n=70), Alpha (n=58), Gamma (n=1), and 20G (n = 39). *p.adj<0.05, **p.adj<0.01, ***p.adj<0.001, ****p.adj<0.0001.

When analyzing only symptomatic samples, we found no statistically significant difference in Ct values amongst the clades (Fig 2C). The benefit of Clemson University’s surveillance strategy is that infections are caught early, often before symptoms are present, which decreases the number of symptomatic samples in our population. While there are significant differences in viral loads between the VOC clades and 20G in pre-symptomatic and asymptomatic patients at the time of initial diagnosis, this trend is not necessarily maintained as the disease progresses. Patients that develop symptoms had higher viral loads regardless of clade. This may explain the apparently contradictory results in the literature; studies which primarily focused on tests from COVID-19 hospitalized patients reported no differences in viral loads among the clades [7], whereas studies that included tests from earlier stage diagnoses reported significant differences in viral loads, particularly for Delta [5-6, 27].

Additionally, patients that report symptoms are much more likely to test positive compared to non-symptomatic patients (Fig 3). From January to November 2021, the average positivity rate for symptomatic samples was 12.71% and for surveillance samples was 0.98%. During the surge in cases due to the Alpha variant in March 2021, samples from patients at the community site who reported exposure were much more likely to be positive for SARS-CoV-2 when compared to non-exposed (8.8% vs 1.7%). However, after the emergence of Delta, the test positivity rate was 10% in both groups. This is likely due to the overwhelming presence of Delta within our community and the extremely high viral load, likely ensuring that everyone had some level of exposure.

**Fig 3:**
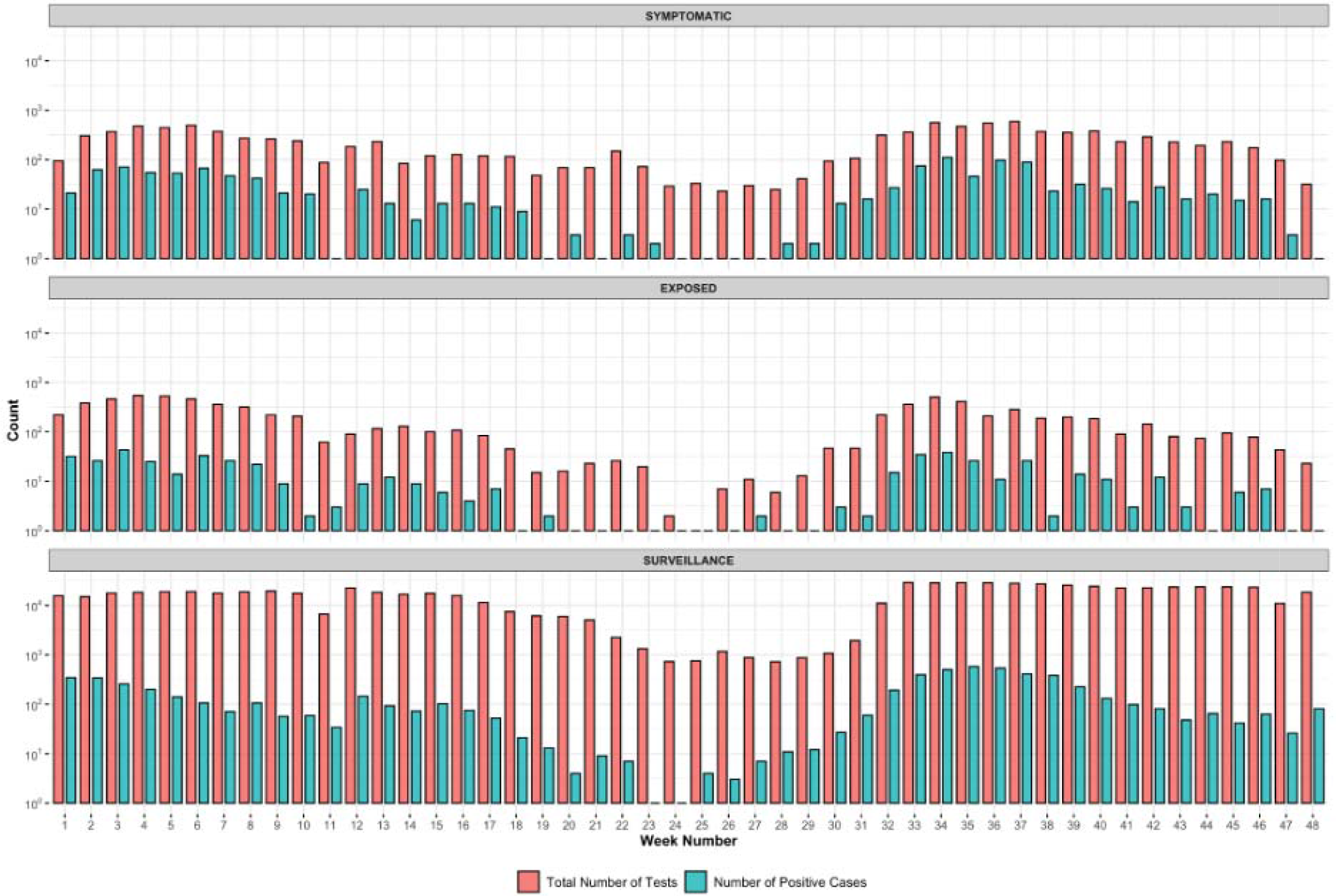
Number of tests and positive tests per category, by week. Note that the y-axis is on a log10 scale. Samples are labeled “symptomatic” if the patient reports symptoms at the time of testing, or labeled “exposed” if they report exposure to a positive patient. Surveillance samples represent the rest of the samples collected. The lower case load during week 11 is due to the university’s spring break, and weeks 18-29 account for summer break.

Due to a non-normal data distribution (skew=-0.307, kurtosis=2.780), we performed Kruskal-Wallis test for stochastic dominance. However, it has been suggested that ANOVA is robust to slight non-normality, such as our data [28-30]. Reanalyzing the data with Welch’s ANOVA, we observed similar results (SFig 2) and determined there was approximately an 8-fold difference in viral load between Delta and 20G and a 2-fold difference between Delta and Alpha, which are consistent with other studies using NP swabs from initial diagnostic samples [5-6, 27]. Our results highlight the significant difference in Ct values between Delta samples and other VOCs.

## Conclusion

Overall, our study showcases the increased viral load of the Delta variant and provides evidence for its rapid global spread. A major benefit to saliva-based testing is the ease of testing; people are more inclined to test frequently. Specifically, our data show that the Delta VOC has the highest viral load in saliva when compared to 20G, even in healthy, young individuals who are pre- or asymptomatic. These individuals are not often captured by other studies as they are not likely to seek out testing; however, they are known to contribute to the rapid spread of COVID-19 [31]. High infectivity of new variants necessitates accurate surveillance. It is expected that future dominant strains, like the newly emerging Omicron, will have viral loads comparable to or greater than Delta to achieve a competitive advantage.

## Supporting information

Data for Figures 1 and 2

Data for Figure 3

Age and Clade Analysis

## Data Availability

All relevant data are within the manuscript and its Supporting Information files. Data analysis scripts can be found at https://github.com/CUGBF/SARS-CoV-2_Ct-vs-Clade.git

## Acknowledgements

The authors thank Clemson University’s administration, medical staff, and the REDDI Lab who helped develop and implement SARS-CoV-2 testing. Thank you to Mayor Robert Halfacre and Dr. Ted Swann for facilitating Clemson community testing. We thank Dr. Vidhya Narayanan, Dr. David Elms, and Russ Nuttall for technical assistance of whole genome sequencing. Thank you to Kaitlyn Williams, Rachel Ham, Sujata Srikanth, and Jeremiah Carpenter for sample preparation. We thank the REDDI Lab clinical director, Dr. Michael Friez, technical supervisor, Dr. Congyue Peng, and clinical supervisors Lauren Cascio and Wael Namouz for maintaining high complexity certification standards (CLIA number 42D2193465). We thank Rachel Ham and Austin Smothers for their critical reading of this manuscript.

## Supporting Information

**SFig 1:**
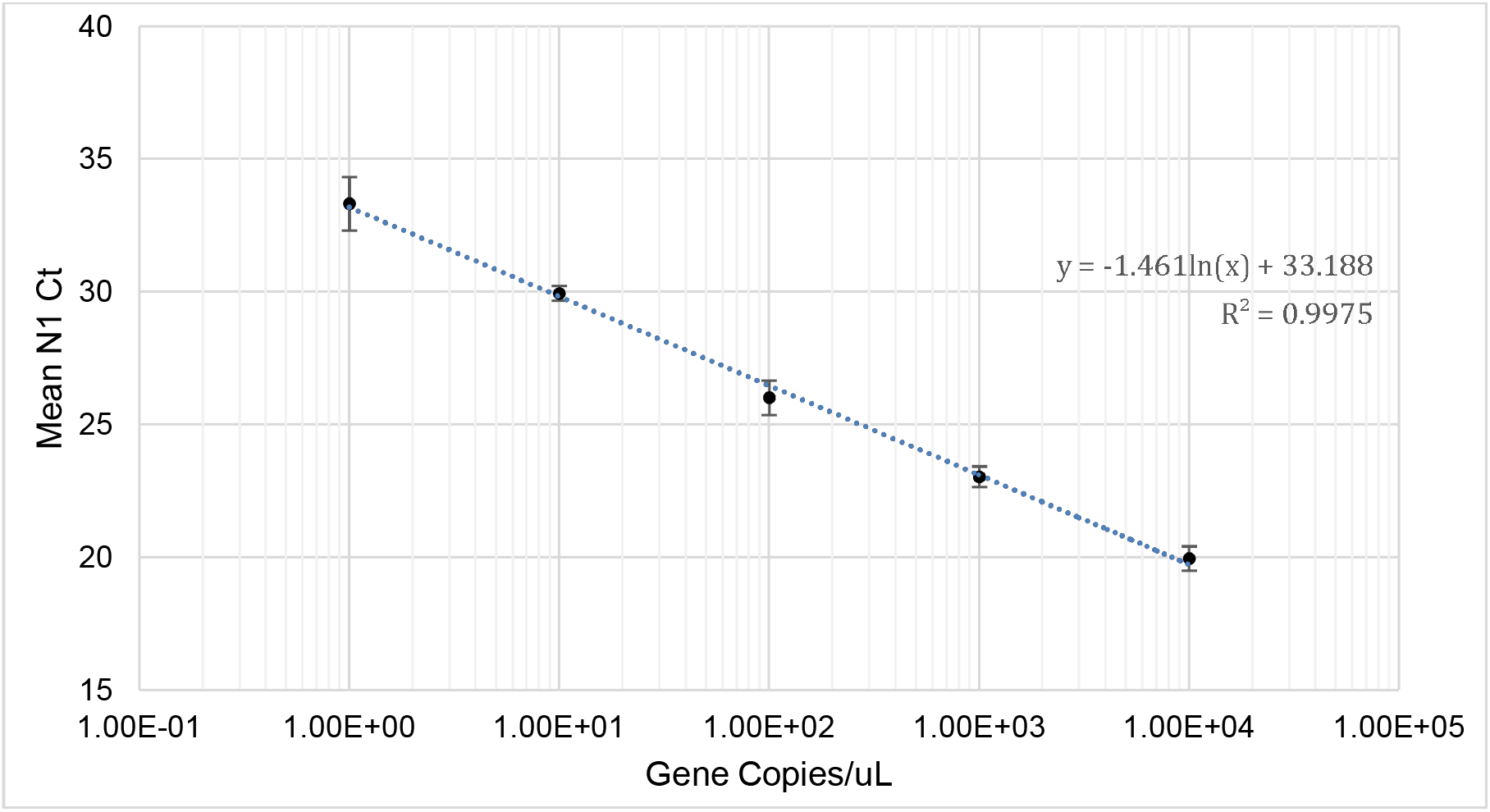
Standard curve for TigerSaliva RT-qPCR assay for N1 detection in synthetic controls. The standard curve was plotted with standard deviations to determine the range of accurate detection using this primer/probe combination. The mean Ct values (n=4) obtained from serial dilutions were plotted against estimated quantify of synthetic RNA in 10μL of RT-qPCR reaction.

**SFig 2:**
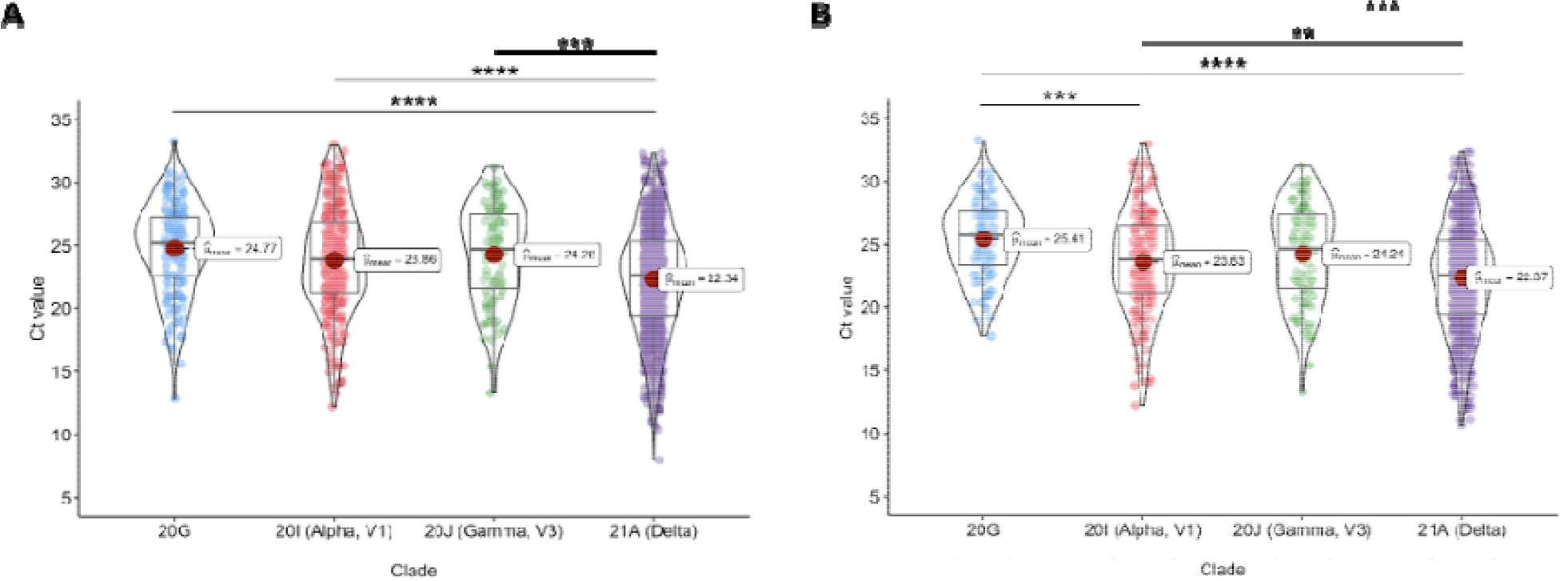
Analysis of Ct values using Welch’s ANOVA test. 2A: Comparison of all samples. We observed a statistically significant difference between Delta and all other clades, including an 8-fold difference in viral load when compared to 20G. **2B: Comparison of only surveillance samples**. The same difference in median Ct was observed between Delta and all other clades. Additionally, surveillance samples showed a statistical difference between Alpha and 20G. *p.adj<0.05, **p.adj<0.01, ***p.adj<0.001, ****p.adj<0.0001

**SFile 1: Accession numbers for sequenced samples uploaded to SCDHEC, GenBank, and GISAID**.

**SFile 2: Demographic Analysis**.

**SFile 3: Data accessibility for Figures 1 and 2**.

**SFile 4: Data accessibility for Figure 3**.

